# Adverse effects of COVID-19 vaccination: machine learning and statistical approach to identify and classify incidences of morbidity and post-vaccination reactogenicity

**DOI:** 10.1101/2021.04.16.21255618

**Authors:** Md. Martuza Ahamad, Sakifa Aktar, Md. Jamal Uddin, Md. Rashed-Al-Mahfuz, AKM Azad, Shahadat Uddin, Salem A. Alyami, Iqbal H. Sarker, Pietro Liò, Julian M.W. Quinn, Mohammad Ali Moni

## Abstract

Good vaccine safety and reliability are essential to prevent infectious disease spread. A small but significant number of apparent adverse reactions to the new COVID-19 vaccines have been reported. Here, we aim to identify possible common causes for such adverse reactions with a view to enabling strategies that reduce patient risk by using patient data to classify and characterise patients those at risk of such reactions. We examined patient medical histories and data documenting post-vaccination effects and outcomes. The data analyses were conducted by different statistical approaches followed by a set of machine learning classification algorithms. In most cases, similar features were significantly associated with poor patient reactions. These included patient prior illnesses, admission to hospitals and SARS-CoV-2 reinfection. The analyses indicated that patient age, gender, allergic history, taking other medications, type-2 diabetes, hypertension and heart disease are the most significant pre-existing factors associated with risk of poor outcome and long duration of hospital treatments, pyrexia, headache, dyspnoea, chills, fatigue, various kind of pain and dizziness are the most significant clinical predictors. The machine learning classifiers using medical history were also able to predict patients most likely to have complication-free vaccination with an accuracy score above 85%. Our study identifies profiles of individuals that may need extra monitoring and care (e.g., vaccination at a location with access to comprehensive clinical support) to reduce negative outcomes through classification approaches. Important classifiers achieving these reactions notably included allergic susceptibility and incidence of heart disease or type-2 diabetes.

## Introduction

The Severe Acute Respiratory Syndrome Coronavirus 2 (SARS-CoV-2) gives rise to COVID-19, the disease causing a massive public health emergency worldwide. This pandemic situation began in December 2019, appearing first in Wuhan, China^1^. This virus is genetically related to certain bat coronaviruses, and its genetic sequence matches 79% and 50% with the coronaviruses responsible for severe acute respiratory syndrome (SARS) and the Middle East respiratory syndrome (MERS)^2^, respectively. The SARS-CoV-2 virus subsequently spread rapidly across the world such that, as of February 2021, and approximately 119 million people have been infected, of which 2.6 million have so far died^3^. This has necessitated strong public health responses and the unprecedentedly rapid development of vaccines against SARS-CoV-2 for public use. These are the first fully validated vaccines designed to combat coronavirus infections in humans^4^, although vaccines for coronaviruses responsible for some non-human diseases have previously been developed^5^. Attempts had previously been made to create vaccines for SARS and MERS, but these so far have been tested only in non-human species^6,7^.

Approximately 66 candidate vaccines for SARS-CoV-2 have been developed, and as of February 2021, of which 17 of these are undergoing phase I trials, 23 are in combined phase I-II trials, 6 in phase II trials, and 20 in phase III trials^8^. Most SARS-CoV-2 vaccines that have completed phase III trials have shown effectiveness in preventing serious disease that is above 90%^8^. Ten vaccines are currently approved by at least one public national health authority for use by their population. These include two RNA vaccines (BNT162b2 from Pfizer–BioNTech and the mRNA-1273 vaccine from Moderna), four conventional inactivated virus vaccines (BBIBP-CorV from Sinopharm, BBV152 from Bharat Biotech, CoronaVac from Sinovac, and WIBP from Sinopharm), three adenoviral vector vaccines (Sputnik V from the Gamaleya Research Institute, the AZD1222 from Oxford–AstraZeneca, and Ad5-nCoV from CanSino Biologics), and a virus peptide fragment-based vaccine EpiVacCorona (from the Vector Institute)^8^.

Numerous countries have begun vaccination programs that prioritise individuals who have the highest probability of severe complications from COVID-19 infection, such as advanced elderly people, and those that are at high risk of virus exposure and transmission such as front-line medical staffs^9^. As of February 2021, over 150 million doses of the COVID-19 vaccine have been administered worldwide^10^. However, no medications are free from adverse reactions. While vaccines protect from many deadly illnesses, all of them may at least demonstrate small episode(s) of significant adverse reactions and side effects, which are evident with their mass administration. This has been documented for COVID-19 vaccines, and it has been generally noted that such affected individuals commonly have pre-existing comorbidities, such as *diabetes and high blood pressure* or allergic conditions.

One issue arising with any vaccine, but particularly in rapidly deployed vaccines is that it can be difficult to appraise the likelihood of adverse reactions to the vaccines. Rarely occurring risk factors are, by the nature and size of the trials and limitations of time, unlikely to be seen in randomized clinical trials. Clinical and demographic information at the individual level can also affect vaccine response. Note that vaccine adverse reactions are a quite separate issue to vaccine effectiveness in preventing disease although they may share some common factors, such as the strength of the immune response to the vaccine. Adverse reactions reported to date are rare, but some uncommon allergic reactions that can develop within minutes to hours after vaccination have been reported as due to the *anaphylaxis*, which can prove very serious for some individuals^20^. Physicians and researchers at the US Centers for Disease Control and Prevention (CDC) assessed adverse reactions after vaccination to identify these reports as *anaphylaxis* or *not anaphylaxis*^21^. In the USA, 1,893,360 people have received their first doses of vaccine since December 14 to 23, 2020^22^, among which 21 cases were reported to involve anaphylaxis responses as identified by CDC; of these 21, 4 were hospitalized and 17 were treated in an emergency department^14^.

Some reported cases of side effects have been noted after SARS-CoV-2 vaccination; in addition there have been a small number of fatalities^27^, although the degree to which they are linked to vaccination is unclear and under investigation^28,30^. Nevertheless, there is a chance of side effects with any medication administered to in a very large population, necessitating close surveillance to detect any evidence of direct or indirect effects. Although the number of cases where adverse reactions to COVID-19 were observed is extremely small in number relative to the number vaccinated, they cannot be overlooked as they give important information to predict and ameliorate adverse reactions and poor outcomes. Statistical and machine learning analysis would play a key role in characterizing and prioritizing those factors. We have, therefore, analysed data from patients to clarify the common causes of such reactions. We employed statistical analysis and trained machine learning models to identify individuals who are most at risk of vaccine complications. If the causes of adverse effects of a vaccine are identified and eliminated, and patients identified as at risk of complications vaccinated in a safe medical environment it would prevent serious conditions developing and enable rapid treatment of anaphylaxis, making COVID-19 vaccination much safer.

The main findings of this study are -

- To identify the most significant features of patient past medical history that can give rise to adverse effects of SARS-CoV-2 vaccination
- To find the most significant patient symptoms that can predict the patient need for hospitalization for treatments after SARS-CoV-2 vaccination
- In cases of death recorded after SARS-CoV-2 vaccination, to find the contributing causes of death
- To identify and classify by the machine learning methods those patients at high medical risk of severe adverse reaction after COVID-19 vaccination and needs of extra precautions

## Results

In this study, we have used two different types of factors with two different analyses and then correlated each of the results. The type of factors employed includes features of the medical history of the patients who demonstrated reactions after vaccination, and the reaction natures were symptoms that arose after vaccination.

### Distribution of patient medical history features and reactions

In this section, we describe the percentage of each significant factor of patient medical history and reactions shown in Table 1. Although the average age of the individuals was about 57 years old, the age of those cases of fatalities and hospitalizations was about 76 and 60 years, respectively. Thus, there is a clear difference in age between different patient groups. Two-thirds of the individuals receiving the vaccines received only the first of an intended two doses. In our study, there were twice the number of female participants compared to male participants and almost half of them were recorded as regularly taking other medications. A history of allergies was a frequently observed factor, with approximately 1 in 4 of the total cases and close to 1 in 3 of the fatality cases. In the hospitalized patient group those with a history of allergies made up 2 in 5 but, in contrast, there were comparatively much fewer among SARS-CoV-2 positive patients. Other common diseases associated with significant patient reactions included *type-2 diabetes, hypertension*, and *heart disease* which together account for around 10% while all the remaining factors cumulatively accounted for 2%-3%.

**Table 1.** The vaccination, demographic and patients’ medical history.

| Patients' Group | All patients'<br>n=5209 | Died<br>n=780 | SARS-CoV-2 Positive<br>n=916 | Hospitalized<br>n=1716 |
| --- | --- | --- | --- | --- |
| Features | count(%) | count(%) | count(%) | count(%) |
| **Vaccination Information** |  |  |  |  |
| **Vaccine Dose** |  |  |  |  |
| Dose 1 | 3418(65.62) | 540(69.23) | 715(78.06) | 1069(62.44) |
| Dose 2 | 864(16.59) | 121(15.51) | 21(2.29) | 366(21.38) |
| Unknown | 927(17.8) | 119(15.26) | 180(19.65) | 277(16.18) |
| Days to Onset (average) | 3.58 | 5.34 | 7.04 | 3.42 |
| **Demographic Information** |  |  |  |  |
| Age (average) | 57.14 | 76.37 | 57.17 | 60.29 |
| **Gender** |  |  |  |  |
| Male | 1631(31.31) | 424(54.36) | 266(29.04) | 660(38.55) |
| Female | 3287(63.1) | 341(43.72) | 484(52.84) | 1032(60.28) |
| Unknown | 291(5.59) | 15(1.92) | 166(18.12) | 20(1.17) |
| **Patients' Medical History** |  |  |  |  |
| Taking Other Medicine | 2405(46.17) | 425(54.49) | 175(19.1) | 1004(58.64) |
| Prior Vaccine | 114(2.19) | 3(0.38) | 4(0.44) | 37(2.16) |
| Allergic History | 1315(25.24) | 231(29.62) | 75(8.19) | 680(39.72) |
| Type-2 Diabetes | 494(9.48) | 156(20) | 47(5.13) | 258(15.07) |
| Hypertension | 707(13.57) | 242(31.03) | 66(7.21) | 369(21.55) |
| Arthritis | 179(3.44) | 47(6.03) | 11(1.2) | 81(4.73) |
| Asthma | 293(5.62) | 17(2.18) | 24(2.62) | 115(6.72) |
| Migraine | 121(2.32) | 1(0.13) | 5(0.55) | 56(3.27) |
| High cholesterol | 116(2.23) | 22(2.82) | 13(1.42) | 33(1.93) |
| Abnormal Blood Pressure | 150(2.88) | 16(2.05) | 15(1.64) | 47(2.75) |
| COPD | 137(2.63) | 56(7.18) | 10(1.09) | 75(4.38) |
| GERD | 181(3.47) | 53(6.79) | 18(1.97) | 92(5.37) |
| Anxiety | 169(3.24) | 41(5.26) | 15(1.64) | 82(4.79) |
| Obesity | 99(1.9) | 26(3.33) | 12(1.31) | 57(3.33) |
| Depression | 171(3.28) | 41(5.26) | 12(1.31) | 75(4.38) |
| Thyroid Disorder | 315(6.05) | 74(9.49) | 32(3.49) | 137(8) |
| Anemia | 117(2.25) | 51(6.54) | 9(0.98) | 63(3.68) |
| Dementia | 138(2.65) | 85(10.9) | 14(1.53) | 55(3.21) |
| Cancer | 107(2.05) | 28(3.59) | 8(0.87) | 56(3.27) |
| Kidney Disease | 205(3.94) | 93(11.92) | 25(2.73) | 106(6.19) |
| Hyperlipidemia | 244(4.68) | 99(12.69) | 30(3.28) | 143(8.35) |
| Heart Disease | 330(6.34) | 156(20) | 26(2.84) | 156(9.11) |
| Covid-19 Positive History | 129(2.48) | 16(2.05) | 14(1.53) | 23(1.34) |
| Atrial Fibrillation | 89(1.71) | 54(6.92) | 7(0.76) | 35(2.04) |
| Pain Symptoms | 111(2.13) | 28(3.59) | 12(1.31) | 42(2.45) |

The reactions of patients are shown in Table 2. It can be seen that *patient disability, headache, pyrexia, dyspnoea, fatigue*, and *chills* count was near 10% of the total cases observed. The next most frequent adverse reactions include *different kinds of pain, dizziness, nausea, asthenia, and vomiting* fall mainly in the range 5% to 10%, with the incidence of other maladies below 5%.

**Table 2.** The adverse reactions after COVID-19 vaccination

| Patients' Group | All patients'<br>n=5209<br>count(%) | Died<br>n=780<br>count(%) | SARS-CoV-2 Positive<br>n=916<br>count(%) | Hospitalized<br>n=1716<br>count(%) |
| --- | --- | --- | --- | --- |
| **Patients' Adverse Reactions** |  |  |  |  |
| Disable | 304(5.84) | 2(0.26) | 4(0.44) | 102(5.96) |
| Headache | 673(12.92) | 21(2.69) | 69(7.53) | 192(11.21) |
| Pyrexia | 664(12.75) | 47(6.03) | 85(9.28) | 283(16.53) |
| Dyspnoea | 555(10.65) | 82(10.51) | 60(6.55) | 306(17.87) |
| Fatigue | 530(10.17) | 39(5) | 61(6.66) | 160(9.35) |
| Chills | 486(9.33) | 11(1.41) | 40(4.37) | 135(7.89) |
| Pain | 468(8.98) | 15(1.92) | 52(5.68) | 138(8.06) |
| Dizziness | 467(8.97) | 14(1.79) | 15(1.64) | 175(10.22) |
| Nausea | 441(8.47) | 29(3.72) | 20(2.18) | 189(11.04) |
| Pain in extremity | 351(6.74) | 10(1.28) | 38(4.15) | 78(4.56) |
| Asthenia | 316(6.07) | 42(5.38) | 36(3.93) | 157(9.17) |
| Vomiting | 271(5.2) | 41(5.26) | 12(1.31) | 155(9.05) |
| Malaise | 256(4.91) | 28(3.59) | 55(6) | 66(3.86) |
| Cough | 248(4.76) | 28(3.59) | 102(11.14) | 85(4.96) |
| Injection site pain | 223(4.28) | 7(0.9) | 7(0.76) | 37(2.16) |
| Myalgia | 219(4.2) | 7(0.9) | 21(2.29) | 64(3.74) |
| Hypoaesthesia | 219(4.2) | 1(0.13) | 0(0) | 93(5.43) |
| Chest pain | 208(3.99) | 16(2.05) | 5(0.55) | 145(8.47) |
| Feeling abnormal | 198(3.8) | 11(1.41) | 30(3.28) | 49(2.86) |
| Rash | 196(3.76) | 4(0.51) | 2(0.22) | 55(3.21) |
| Condition aggravated | 192(3.69) | 25(3.21) | 12(1.31) | 83(4.85) |
| Chest discomfort | 186(3.57) | 5(0.64) | 5(0.55) | 76(4.44) |
| Arthralgia | 186(3.57) | 5(0.64) | 8(0.87) | 37(2.16) |
| Paraesthesia | 184(3.53) | 0(0) | 3(0.33) | 73(4.26) |

| **Patients' Group** | **All patients'** | **Died** | **SARS-CoV-2 Positive** | **Hospitalized** |
| --- | --- | --- | --- | --- |
| Unresponsive to stimuli | 168(3.23) | 107(13.72) | 11(1.2) | 63(3.68) |
| Diarrhoea | 167(3.21) | 16(2.05) | 22(2.4) | 74(4.32) |
| Pruritus | 148(2.84) | 2(0.26) | 2(0.22) | 31(1.81) |
| Heart rate increased | 148(2.84) | 3(0.38) | 3(0.33) | 52(3.04) |
| Urticaria | 146(2.8) | 0(0) | 0(0) | 39(2.28) |
| Facial paralysis | 143(2.75) | 0(0) | 0(0) | 41(2.39) |
| Syncope | 139(2.67) | 29(3.72) | 1(0.11) | 58(3.39) |
| Tachycardia | 136(2.61) | 8(1.03) | 5(0.55) | 58(3.39) |
| Palpitations | 134(2.57) | 5(0.64) | 3(0.33) | 58(3.39) |
| Hyperhidrosis | 128(2.46) | 5(0.64) | 5(0.55) | 47(2.75) |
| Erythema | 125(2.4) | 8(1.03) | 0(0) | 27(1.58) |
| Throat tightness | 124(2.38) | 0(0) | 0(0) | 44(2.57) |
| Tremor | 121(2.32) | 7(0.9) | 3(0.33) | 46(2.69) |
| Blood pressure increased | 120(2.3) | 4(0.51) | 2(0.22) | 46(2.69) |
| Anaphylactic reaction | 112(2.15) | 2(0.26) | 0(0) | 40(2.34) |
| Intensive care | 113(2.17) | 16(2.05) | 8(0.87) | 103(6.02) |
| Loss of consciousness | 106(2.03) | 10(1.28) | 0(0) | 36(2.1) |
| Decreased appetite | 103(1.98) | 32(4.1) | 16(1.75) | 37(2.16) |
| Muscular weakness | 97(1.86) | 2(0.26) | 3(0.33) | 51(2.98) |
| Flushing | 100(1.92) | 0(0) | 3(0.33) | 26(1.52) |
| Mobility decreased | 98(1.88) | 6(0.77) | 4(0.44) | 43(2.51) |
| Injection site erythema | 97(1.86) | 0(0) | 0(0) | 15(0.88) |
| Feeling hot | 96(1.84) | 2(0.26) | 3(0.33) | 28(1.64) |
| Abdominal pain | 95(1.82) | 8(1.03) | 0(0) | 64(3.74) |
| Injection site swelling | 93(1.79) | 0(0) | 1(0.11) | 15(0.88) |
| Cerebrovascular accident | 91(1.75) | 17(2.18) | 1(0.11) | 73(4.26) |
| Cardiac arrest | 92(1.77) | 76(9.74) | 4(0.44) | 22(1.29) |
| Lymphadenopathy | 91(1.75) | 0(0) | 3(0.33) | 17(0.99) |
| Died | 780(14.97) | 780(100) | 69(7.53) | 116(6.78) |
| SARS-CoV-2 test positive | 916(17.58) | 69(8.85) | 916(100) | 120(7.01) |
| Hospitalized | 1712(32.87) | 116(14.87) | 120(13.1) | 1712(100) |

### Finding significant and associations between patient medical history factors and post-vaccination adverse reactions using statistical analyses

Using two different statistical tests, we identified the most associative and significant parameters including patients’ medical history factors (including pre-existing diseases and other discomforts) and identified the adverse reactions or symptoms that may have predisposed to the development of severe health conditions, even fatality. In this analysis, we considered those significant parameters with a value of P < 0.05, or lower. The target variables that we have used in our statistical analyses were, *death, SARS-CoV-2 positive status*, and *hospital admission status*.

In term of patients’ medical histories, *age, gender, type-2 diabetes, allergic history and hypertension* were the most significant features among all the target groups. But for patients’ death status *anemia, atrial fibrillation, dementia*, and *heart disease* were also found as significant. Some of the significant parameters are that are common for hospitalised and SARS-CoV-2 positive patients were *allergic history, anemia, hyperlipidemia* and *kidney disease*. However, the *high blood pressure*, and *high cholesterol* were not found as significant within any of the groups. The details results for this analysis are shown in Figure 2.

**Figure 1.**
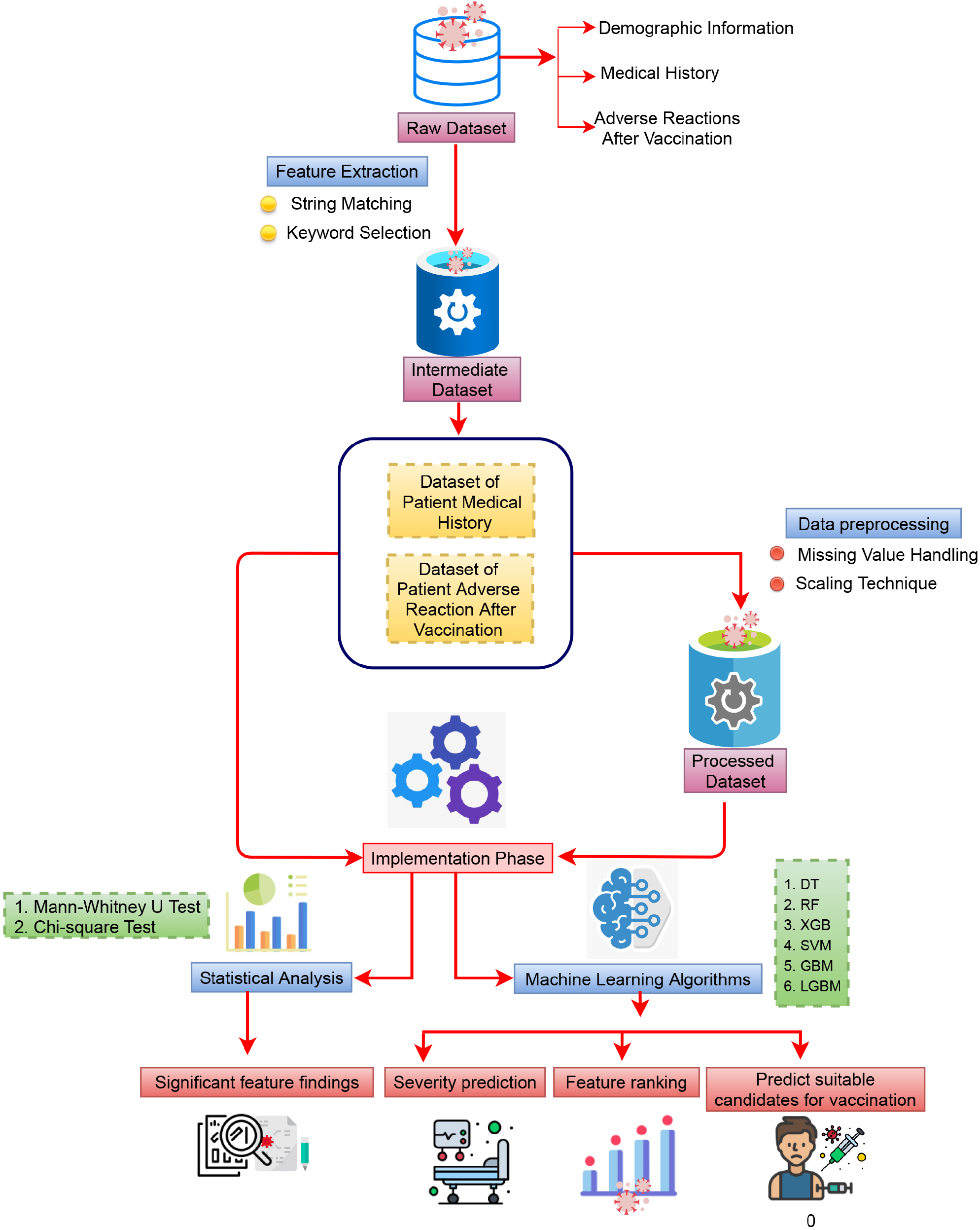
The schematic diagram of the overall workflow including data processing, data division, analysis using statistical and machine learning methods, and at the end performance evaluation with finding significant features.

**Figure 2.**
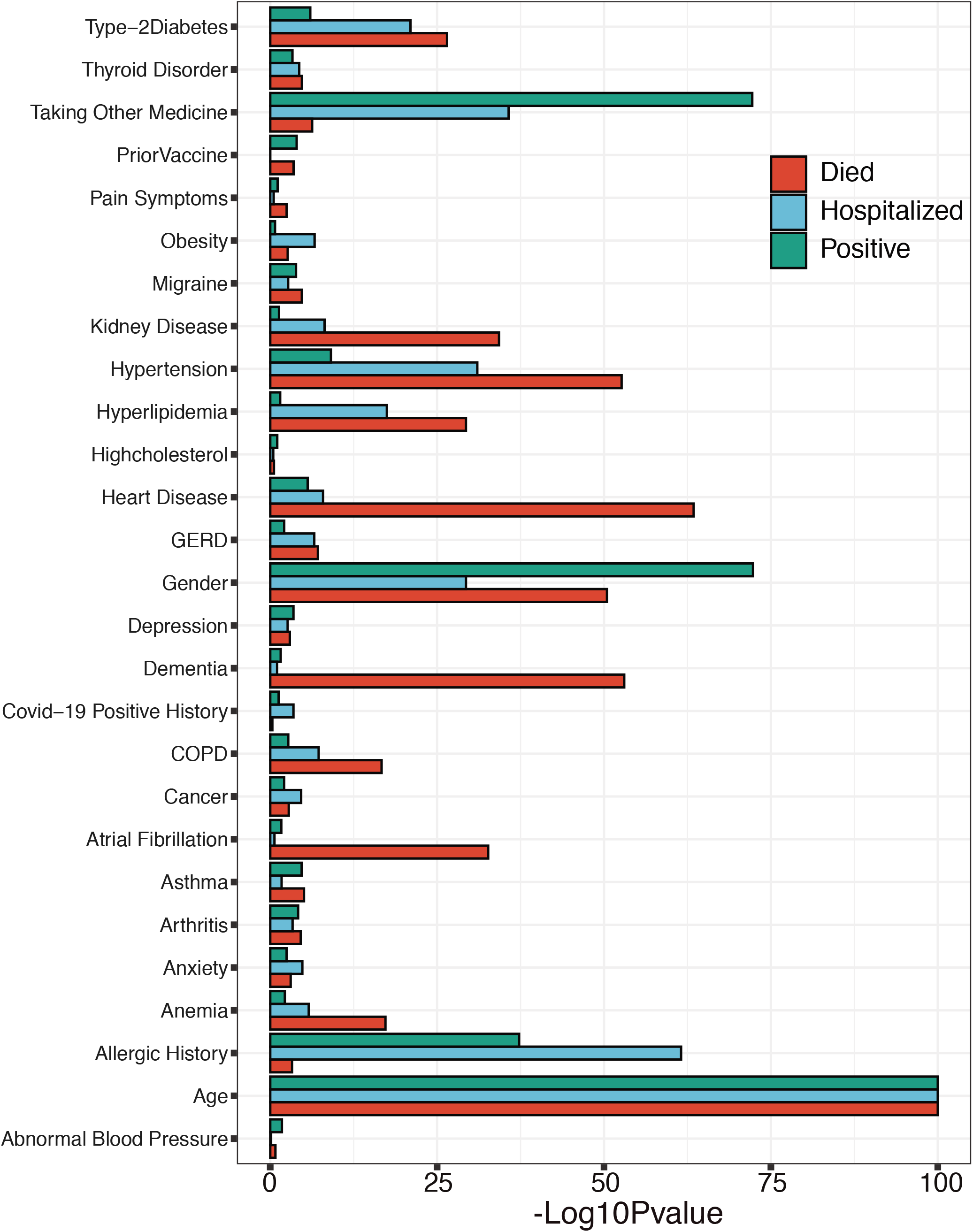
The significant features within the patients’ medical history, where the higher bar length indicates greater the

We have also performed a similar analysis for the dataset with patient adverse reactions, and identified a list of significantly associated symptoms that are shown in Figure 3. In this case, we also considered three target variables as independent variable, when it is not considered as a target variable on the time of analysis. It can be observed that the *patients’ died status, hospital stay duration in days, hospitalized status*, and *SARS-CoV-2 positive status* were the common factor for all the three target variables. When we have considered the incidence of patient mortality as a target variable, *cardiac arrest, unresponsive stimuli*, and *different kinds of pain* were found to be the most significant. It was also be observed that *abdominal pain, cerebrovascular pain, dyspnoea, intensive care treatment, and vomiting* were found as significant for the hospitalization status, whereas the *cough, nausea, and syncope* were for SARS-CoV-2 positive status. In addition to that, the *chest pain, chills, patients’ disability, dizziness, facial paralysis*, and *hypoaesthesia* were commonly found as a significant factor for patients’ death and SARS-CoV positive status.

**Figure 3.**
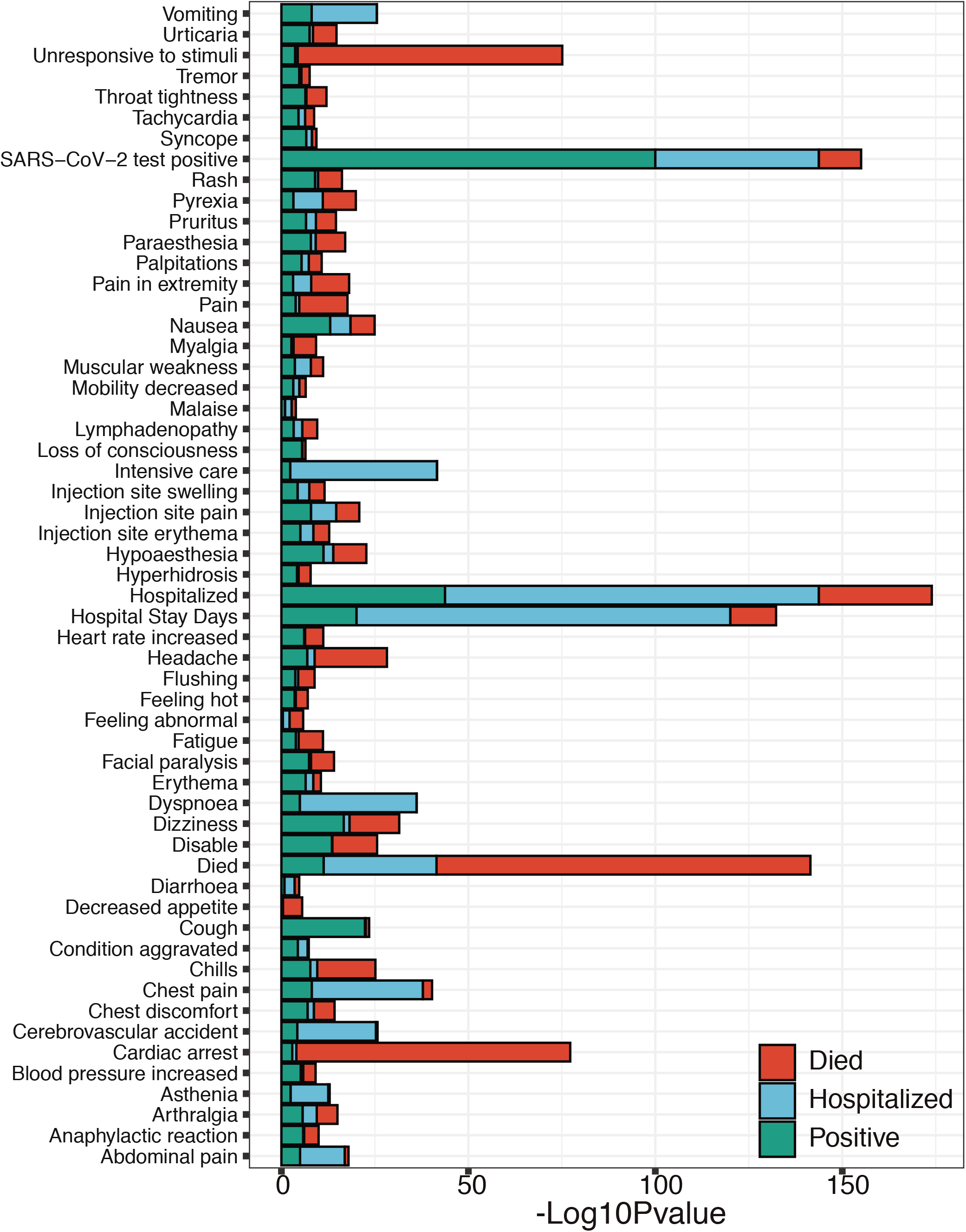
The significant patients’ adverse reactions after vaccination, where the higher bar length indicates greater the

### Feature Importance analysis for finding significant features using machine learning classifiers

After model training, we calculated the coefficient values for each of the features and prioritized them as significant with regards to their corresponding target variables. Firstly, we calculated the feature importance scores for each distinct feature, for individual machine learning classifiers, and then we normalized the values to render the data with the same scale, i.e. between 0 and 1, by using the min-max normalization technique^13^. This was followed by the mean aggregation of those values as shown in Figure 6.

In the case of patient past medical histories, the identified features are shown in Figure 6.A, where the patient *age, gender, taking other medicine, heart disease, allergic history, hypertension* and *type-2 diabetes* have shown significant importance when the target variable was the patients’ *death status*. With the target variable *SARS-CoV-2 positive status*, it is also shown that other than the *hyperlipidemia*, similar features were found as significant as with the results for *death status*. Lastly, for the *hospital admission status* as a target variable, we have found similar features are common with the other two groups.

Figure 6.B shows the importance of features listed according to the category of patient post-vaccination adverse reactions or symptoms. For the first target variable (i.e., *patient mortality status*), the most important features identified were the *hospital treatment duration, unresponsive stimuli, dizziness, headache, physical disability, cardiac arrest, chills, rash, facial paralysis* and *various kind of pain*. For the second target variable (i.e., SARS-CoV-2 test status), the significant features were similar to the case of the first target variable including that the *dyspnoea* was a novel finding as an important factor. Finally, for the third target variable, *hospital admission*, the significant features identified were *hospital treatment duration, pyrexia, fatigue, headache, dizziness, rash* and *dyspnoea*.

### Classification of Patients using Machine Learning algorithms

In our machine learning analysis, firstly we considered the patient medical histories as the independent features, and the *patient death, SARS-CoV-2 test positive*, and *hospital admission status* as dependent features, which depends on those independent features. Next, by considering both patient medical history and patient reactions after vaccination. First, we trained our models and evaluated their performances with the test data by calculating a range of metrics including *accuracy, precision, recall, F1-score, ROC-AUC* and *Log-loss*, which are shown in Figure 4.A including the ROC-AUC curves which is shown in the Figure 5 in the panel A, B and C, respectively for patients medical history. The results show that, when the target feature variable was the patient death status, then the RF and XGB models performed the best among all with an accuracy of 0.89, precision scores of 0.90 and 0.89, recall scores of 0.88, F1-score of 0.90, AUC score of 0.89, and the log-loss of 3.76% and 3.90%, respectively. However, excluding SVM, other model performances were also encouraging when the scores of similar performance metrics were observed. Similar observations were made when we considered *the SARS-CoV-2 test result* as the target feature variable, i.e. the XGB outperformed other competing methods, with 0.86 accuracy scores, respectively, while other models performances were also found as competitive except SVM, which achieved almost consistently below 0.80, and the log-loss were also higher than others (i.e. above 10%). Finally, for the target variable, *hospital admission status*, all the models performed almost equally, but score-wise they have demonstrated some performances that are below optimal (i.e. compared to the previous two scenarios).

**Figure 4.**
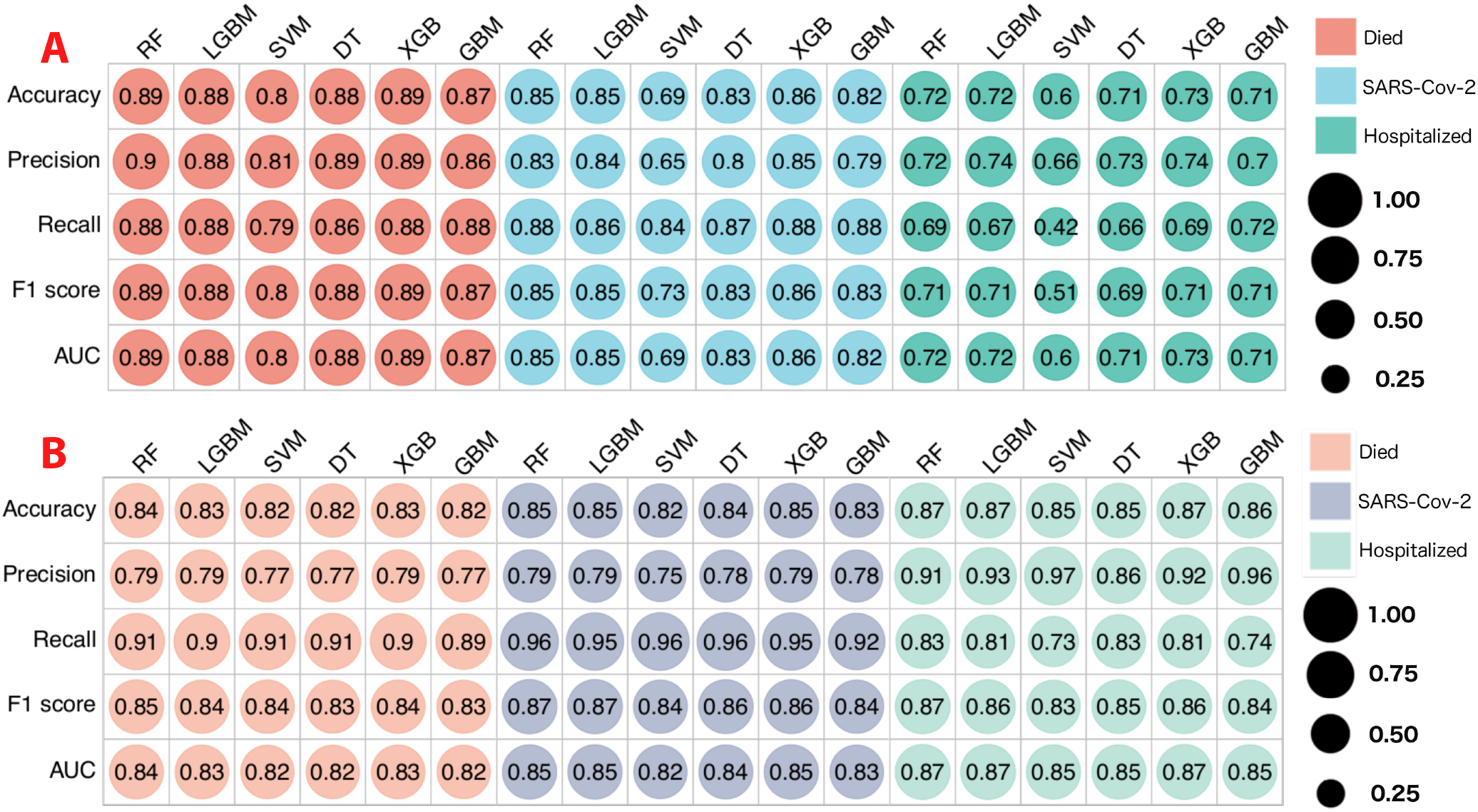
Comparative performance evaluation for the patient classification based on machine learning algorithms; A. with the dataset of patients’ medical history; B. with the dataset of patients’ adverse reactions after vaccination.

**Figure 5.**
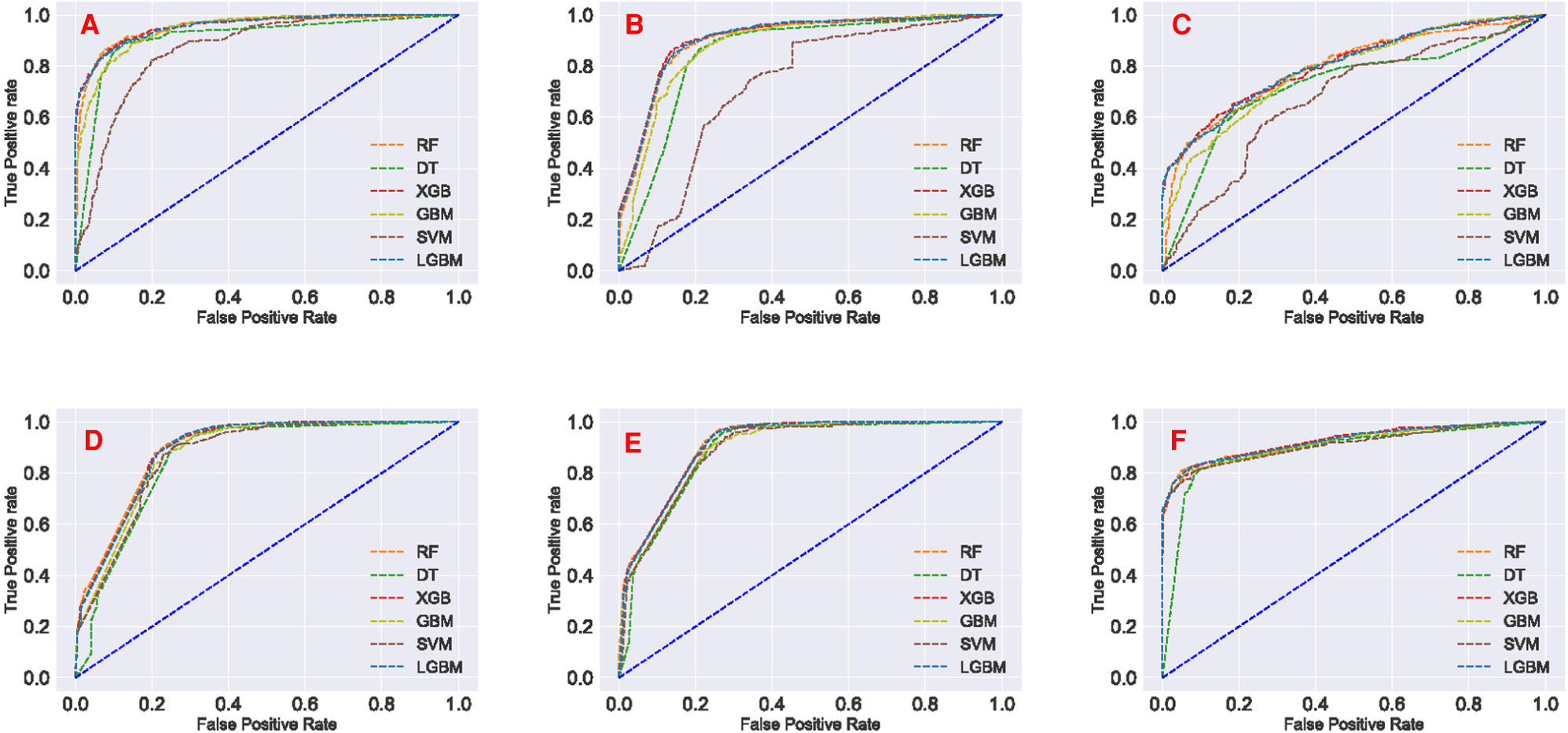
Area Under the ROC curves for the machine learning model evaluation. A. classification of died patients’ using patients’ medical history dataset; B. classification of SARS-CoV-2 positive patients’ using patients’ medical history dataset; C. classification of hospitalised patients’ using patients’ medical history dataset; D. classification of died patients’ using patients’ reaction dataset; E. classification of SARS-CoV-2 positive patients’ using patients’ reaction dataset; F. classification of hospitalised patients’ using patients’ reaction dataset.

**Figure 6.**
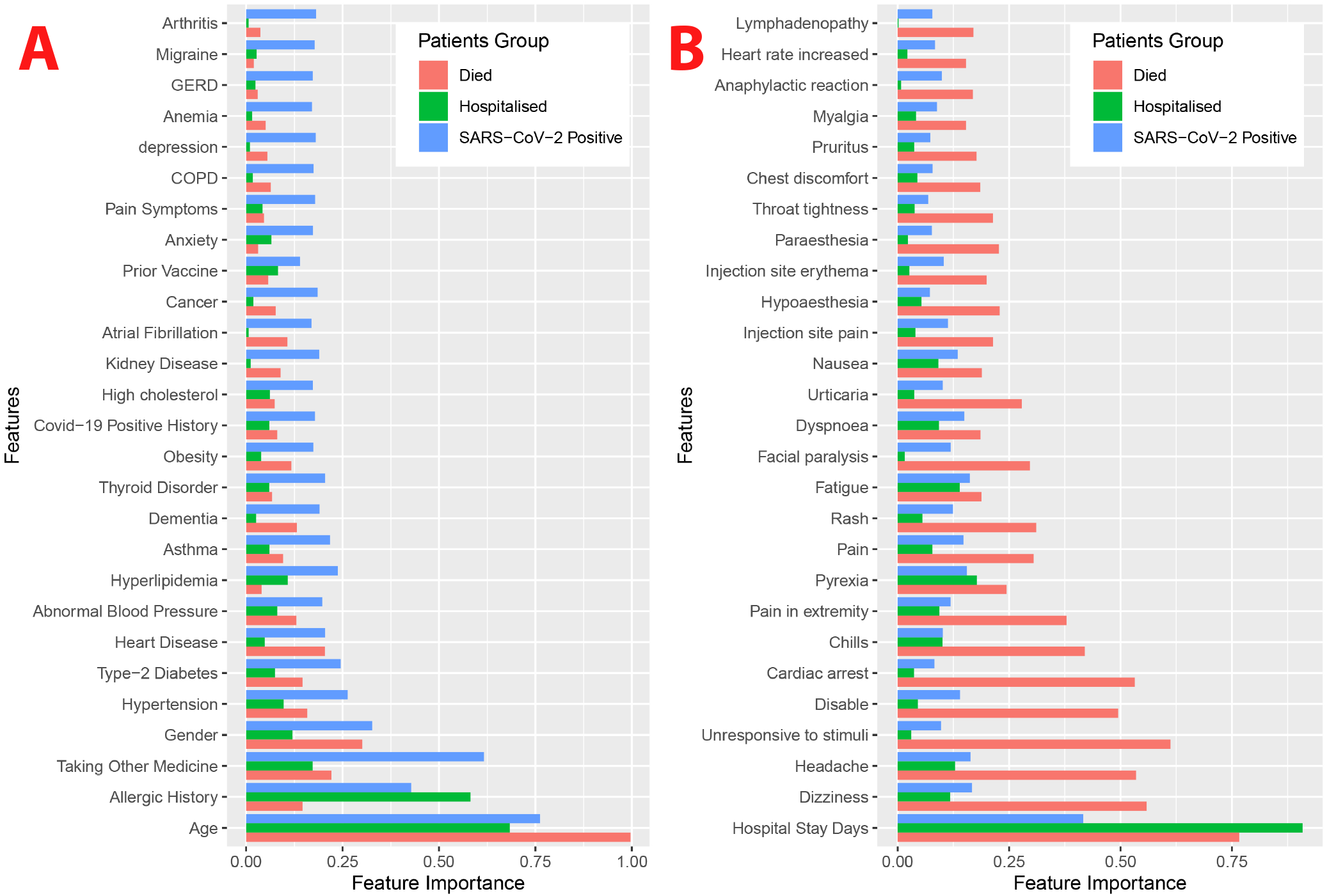
The features ranking according to the coefficient values of the A. Patients’ medical history, B. Patients’ adverse reactions, calculated after machine learning model training. ML model outcomes indicate that higher coefficient values are mostly close to the significant association of severity.

Next, we considered patient post-vaccination adverse reactions as the independent feature and the target variables remain the same as previous. The results indicating model predictive performances are shown in Figure 4.B including the ROC-AUC curves which is shown in Figure 5 in the panel D, E and F, respectively. It can be noted that all the classifiers demonstrated substantially similar performances with scores of greater than 0.80 in all the evaluation matrices and the log-loss was less than 7%. However, it can be also observed that when different target variables were set for the classification tasks after training with the patient adverse reaction, the best performing classifiers (in terms of Accuracy) were different as well, i.e., for the patient *death status*, the RF yielded a score of 0.84. Moreover, for the *SARS-CoV-2 test status*, and *hospital admission status*, the RF, LGBM and XGB yielded 0.84 equally.

## Discussion

Vaccination is a well-accepted reliable approach to prevent diseases^17^, and historically, it has proven to be one of the most effective strategies to control epidemics and pandemics, such as the SARS-CoV-2 outbreak^18^. All vaccines result in at least a small number of patients that demonstrate some kind of post-vaccination side effects^19^. Although vaccines are a life-saving medication, in some cases they can result in an after-effect, and sometimes even resulting in severe symptoms, although with a low probability. It is a challenging task to identify small groups of patients who shows post-vaccination adverse reactions. It is been observed that some patients showing a rapid onset reaction^28^ have needed treatment at a hospital clinic, and in some rare cases, the patient subsequently died^30^.

The main purpose of this research was to determine the key indications that indicate a susceptibility to adverse COVID-19 vaccination effects as well as to identify the key symptoms that indicate the cause or causes of the adverse conditions, including classifying a patient as at high risk or unsuitable for COVID-19 vaccination. We have found a list of the most significant features that support our hypothesis and all of which are commonly found in all the target groups. The most significant demographic information are patients’ *age* and *gender*; most associative patients’ coexisting conditions are *allergic history, taking other medicine, type-2 diabetes, hypertension, and heart disease*; and most significant and associative patients’ after effect of post-vaccination are *duration of hospital treatment, pyrexia, headache, dyspnoea, chills, fatigue, different kind of pain, physical disability, cardiac arrest* and *dizziness*.

Furthermore, some post-vaccination symptoms are commonly found for anonymous cases but a few of them are highly responsible for patients’ severe condition due to vaccination. The most severe side effects identified are the *hospital treatment duration, pyrexia, headache, dyspnoea, chills, fatigue, different kind of pain*, and *dizziness*.

Patient allergic history is commonly associated cause of adverse effects to many types of drugs and vaccines^13^, and in the case of COVID-19 this has also been reported^16,24^; allergic-related reactions are found to a significant degree in every data group used in our study. Patient age is another important aspect, where the mortality rate in advanced old-age persons is comparatively higher than in other cases, and previous findings also supported this evidence^15^. There are reports that indicate allergic history may be a significant issue for COVID-19 vaccination^25,26^. However, our study indicates that patients taking significantly immunosuppressing medications^32^ are at elevated risk of adverse reactions, as are those who are SARS-CoV-2 positive. Our research also suggests that other pre-existing conditions such as *type-2 diabetes, hypertension*, and *heart disease* and a history of allergic responses could also be associated with the development of severe vaccine reactions. Our study also identified a range of other factors linked to significant patient reactions that require hospital treatments and may also be associated with patient mortality.

The utilization of machine learning models is widely acknowledged as capable of demonstrating morbidity/mortality-associated factor identification and for using those factors in making patient outcome predictions^29^. The capacity of machine learning models to find embedded patterns in information by taking into account a large number of factors at once can give an improved understanding of the complex components that underlie phenomena such as vaccination adverse effects. As previously mentioned these machine learning models performs well with our datasets and identified significant parameters related to vaccination-associated symptoms. The models achieved a good accuracy score including good precision, recall, and f1 score as well as low log loss values indicate strong classification and decision making. In our analysis, we saw that several models performed particularly well with high scores for evaluation matrices, i.e., in the dataset of medical history, for the mortality cases 0.89 accuracies for RF and XGB, for the SARS-CoV-2 positive patients 0.86 for XGB, and in the dataset of adverse effects, for the mortality cases RF 0.84, for the SARS-CoV-2 positive and hospitalized patients, RF, LGBM and XGB 0.85 and 0.87 respectively. Thus, based on the exhaustive comparison of various factors utilizing supervised machine learning models, this analysis may identify significant factors to clinicians concerning parameters valuable for patient stratification. In sum, the use of machine learning models such as those presented here to assess the likelihood that a patient is at risk of developing a severe reaction post-vaccination could be of great utility.

Post-vaccination adverse effects could be decreased if at-risk individuals can be identified, which is clearly possible based on the patient medical history, which this study confirms by experimenting with a set of validation datasets. Though vaccination is not directly responsible for patient severe illness or death, we may need very careful observation of identified at-risk patients including access to ICU facilities.

## Conclusion

The results indicate that patient medical histories are strongly related to the incidence of patient adverse reactions, some of which are associated with severe disease and even death. Moreover, a set of significant side effects are also developed as post-vaccination symptoms. Therefore, it is important to identify possible causes of the adverse effects. If recognised, the factors identified can be taken into account by clinicians, enable care improvement.

Based on our analyses, those patients at greatest risk of adverse reaction after vaccination include those of advanced old age, those having allergic conditions, effects, those taking other medications (notably immunosuppressive medications) and those with a history of type-2 diabetes, hypertension or heart disease disorders. Moreover, the study also revealed that a set of symptoms post-vaccination like *hospital stay duration, pyrexia, headache, dyspnoea, chills, fatigue, different kind of pain and dizziness, rash*, and *physical disability* are most associated with severe reactions.

Using statistical and machine learning analysis^31^, we have found factors in patient medical histories that are associated with a risk or adverse patient reaction occurring in the post-vaccination period. Our results also suggest that a common group of severe after-effects, that were identified by the independent analyses, proves that these outcomes are reliable.

Although our analysis reveals significant findings regarding the risk of COVID-19 vaccination effects, there are few limitations that need further research effort. We have used a comparatively small amount of patient data collected from a specific region of the USA, with the mRNA-based vaccines only. Therefore, for making a generalised decision, it is important to have a rigorous analysis with a larger population size. Nevertheless, we hope that the result of this research will play a significant role for policymakers deliberating the distribution of vaccines as well as identifying patients who may be vulnerable to adverse reactions.

The efficacy and safety of COVID-19 vaccines to date are very promising, but minor after-effects of an administered vaccine might be expected and some extreme allergic or other responses may infrequently happen. Although a possibility of post-vaccination adverse effects is not always a reason to avoid vaccines (especially given the serious consequences of COVID-19 in many vulnerable groups), new information about adverse reaction risk that our study provides could be an important consideration in clinical considerations about whether to administer a COVID-19 vaccine and in determining a need for extra monitoring and care at the point of vaccination.

## Methods

In this study, we have considered COVID-19 vaccinated patients data including their past medical history, and their post-vaccination effect and outcomes, and conducted data analyses by applying statistical methods and machine learning models. We also quantify the feature importance values to rank the features after model training.

### Data Collection

In this study, initially we have used a raw dataset of vaccinated USA patients that contains various kind of vaccine-related information. The dataset was collected from the Vaccine Adverse Event Reporting System (VAERS) on 5,351 individuals vaccinated from December 2020 to February 14, 2021, who had also reported adverse reactions^11^. The dataset contains information including COVID-19 vaccination status and the reactions as different sicknesses after vaccination, however, any non-COVID-19 information were omitted from our current study. In this dataset, for the most frequently used mRNA COVID-19 vaccines, the total number of collected reports was 5,209. VAERS collected the patient information on *age, gender, comorbidity history, allergic history, and birth defect information after vaccination, vaccination date, date of reaction onset, hospitalization information after onset, death event, recovery status, and laboratory test information after onset*. All this information was included in the dataset obtained from VAERS and also used in this study.

### Data Processing

Before applying statistical methods and machine learning models, we have pre-processed the dataset, including the use of feature extraction and feature engineering. Applying string matching and keyword selection techniques, we have extracted the patient medical history such as pre-existing non-communicable and communicable diseases, which included *hypertension, diabetes, COPD, kidney disease, depression and asthma*. (detail is shown in Table 1). We have also included the reported adverse reactions including the types of symptoms and signs such as *cough, high temperature, fatigue, fever, pyrexia, nausea, facial paralysis, vomiting* (detail shown in Table 2). We thus obtained a processed dataset with 86 attributes and 5,209 entities.

In the data processing step, especially in feature extraction, we have considered some factors that are described below. Initially, we have extracted and transformed values from the raw textual dataset^11^, i.e. in the “gender” field there were three types of values, i.e. ‘M’ as male, ‘F’ as female, and ‘U’ as unknown gender. In the ‘died’ and ‘disabled’ fields, we have considered ‘Y’ as yes and the remainder are ‘no’; in the ‘prior vaccine’ fields, mentioned vaccine name as ‘yes’ and rests are ‘no’. In the ‘allergic history’ field, we have considered mentioned allergic effects as a positive case of allergic history and the null values, values with ‘no’, ‘none’, ‘NA’, ‘no known allergic effects’ and also more negatively mentioned text as a negative case. But in the ‘History’ column in the raw dataset, coexisting conditions of patients was in written form, we extracted all of the patients’ medical history separately. In this case, we have selected the keywords for each of the features and then matched with the text and found the appropriate medical history, which we have considered as the most frequent top 27 individual medical histories. In the raw dataset^11^, there was a separate file that contains the patents’ adverse reactions as symptoms including a key of ‘VAERS ID’, where we have separated each of the 56 most frequent reactions. There were three different files that were included in the dataset: the first one was for patients’ demographic and medical history, the second one was for patients’ reactions, and the final one was for vaccine information. We have merged the dataset according to the primary key ‘VAERS ID’. Finally, we have eliminated all of the non-COVID-19 vaccinated patients’ data.

We have partitioned our dataset into two different parts. The first part contained the patient medical history and the second part consisted of patient adverse reactions after vaccination (detail of the workflow is shown in Figure 1). After vaccination, some patients died shortly after developing some symptoms, some were re-infected with COVID-19, and some had shown sufficiently severe adverse reactions to require admission to hospital facilities for treatment. For this reason, we consider the three different types of target variables for patient comorbidities and reaction analysis after vaccination. The first one is “death status”, the second one is “SARS-CoV-2 test status” and the third one is patient “hospital admission status” - all of which were observed after vaccination.

Furthermore, for the machine learning algorithms, we have performed some additional steps to process the data. For the data field, namely *the age*, approximately 19.22% of data was missing, which were imputed with the mean value. Before each of the *train-test split* of the dataset, we have standardized our dataset with zero mean and unit standard deviation^23^.

Among all of the 5,209 COVID-19 vaccinated individuals, 780 have died, 916 were re-infected to COVID-19, and 1712 were admitted to hospital for treatment due to serious post-vaccination illness. Next, we considered those attributes as independent variables and perform statistical and machine learning analysis.

### Statistical and Machine Learning Approaches

We have used statistical and machine learning approaches to find the significant features; and machine learning models also capable of distinguishing between the various group of patients’. For the categorical variables, we had used the chi-square test for finding the corresponding P values and considers P < 0.05 as a significant as well as associative parameter. Because age is absolute discrete data, we have used the Mann Whitney U test over two different populations. We had also performed descriptive statistical analysis to calculate the percentage and mean values of the features. In machine learning analysis, there was a range of models i.e, decision tree (DT) and random forest (RF) are tree-based algorithms, support vector machine (SVM) is kernel-based and three boosting algorithm, gradient boosting machine (GBM), extreme gradient boosting machine (XGB) and light gradient boosting machine (LGBM)^31^. We selected those supervised Machine learning algorithms for classification because of their excellent performance and quick execution^33^. For this purpose, classifiers that are based on max voting, averaging and weighted-averaging have used as a basic ensemble learning approach, along with that, the advance ensemble learning approach also functions as stacking, bagging and boosting. Those techniques are highly efficient and easy to debug^34^.

In the model training phase, the machine learning algorithms had some parameters to extract significant features. In the Decision tree algorithm, the random state set as 42 with minimum sample split number two and ‘gini’ used as a criterion. Random forest was used as same as a Decision tree with a minimum of two split samples. On the other hand, SVM sets as a linear kernel. The learning rate was 0.1 with criterion ‘friedman_mse’ in GBM. But the learning rate of LGBM was 0.05 with a bagging fraction 0.8 and a bagging frequency 5. A tree-based booster with max depth six was used in the XGB algorithm and the learning rate was 0.1.

To evaluate the machine learning models, a set of matrices are used i.e. accuracy, precision, recall, f1-score, AUC, and log losses. To find the associative parameters we calculate the feature importance values for every machine learning model. The coefficient values of each feature represent the corresponding contribution of model training to separate an unknown instance among classes.

## Data Availability

The dataset and corresponding codes are available on the following repositories:
Dataset: https://vaers.hhs.gov/data/datasets.html
Codes: https://github.com/m-moni/COVID-1

## Author contributions

The research presented in this article is a combined effort of eleven authors. The article is prepared by executing several phases of the research. At the early stage of the research,Md. Martuza Ahamad (MMA) and Sakifa Aktar (SA) had collected the data from VAERS repository on COVID-19 vaccination information according to the direction of Mohammad Ali Moni (MAM). Next, the four authors joining with Md Jamal Uddin (MJU) designed the architecture of the workflow. The other authors are Md. Rashed-Al-Mahfuz (MRM), AKM Azad (AKMA), Shahadat Uddin (SU), Salem A. Alyami (SAA), Iqbal H. Sarker (HIS), Pietro Liò (PL), Julian M.W. Quinn (JMWQ). Based on that workflow, the authors contributed as follows:

MMA, SA, MJU and MMA: They collected vaccinated patient’s data from the dataset and make them fit for our experiments. They conducted most of the experiments and joined every meeting of the research discussion. They took part in writing the primary draft of the article as well as group-wise reviewing the article at overleaf.

MRM, AKMA, SU, SAA, HIS, PL and JMWQ:They were involved in the writing and reviewing the whole article.MAM:He supervised the whole work. Additionally, he conducted several experiments and took part in every writing phase of the article.

MMA and SA act as a principle investigator of this work in contributed equally.

## Competing interests

The author(s) declare no competing interests.

## Data and Code availability

The dataset and corresponding codes are available on the following repositories:

- Dataset: https://vaers.hhs.gov/data/datasets.html
- Codes: https://github.com/m-moni/COVID-19

## Consent and Ethics Approval

We have used publicly available anonymous data (https://vaers.hhs.gov/data/datasets.html) in this study. Therefore, we do not need any kind of approval for experimental protocols, consents, guidelines and regulations for this study.

## Notes

### Competing Interest Statement

The authors have declared no competing interest.

### Funding Statement

Author(s) don't get any types of funds for this research.

### Author Declarations

This work considers the publicly available dataset, that's why there is no need to get any kinds of approvals.

